# Assessing Patient Satisfaction in Ibadan South-west Region of Oyo State, Nigeria

**DOI:** 10.1101/2024.03.04.24303744

**Authors:** Abel Nnandi Chukwuemeka, Folahanmi Tomiwa Akinsolu, Marylyn Bukola Adekola, Mobolaji Timothy Olagunju, Olunike Rebecca Abodunrin, Ifeoluwa Eunice Adewole, Oluwabukola Mary Ola, Oliver Chukwujekwu Ezechi

**Affiliations:** Department of Public Health, Faculty of Basic Medical and Health Sciences, Lead City University Ibadan, Oyo State, Nigeria; Department of Public Health, Southern Illinois University Edwardsville, USA; Department of Epidemiology and Biostatistics, Nanjing Medical University, China; Clinical Science Department, Nigerian Institute of Medical Research, Lagos, Nigeria; Center for Reproduction and Population Health Studies, Nigerian Institute of Medical Research, Lagos, Nigeria; Department of Planning, Research, Monitoring and Evaluation, Lagos State Health Management Agency, Alausa, Lagos. Nigeria

**Keywords:** Patient Satisfaction, Healthcare Facilities, Percentage of Positive Responses (PPR), Short-form Patient Satisfaction Questionnaire (PSQ-18), Ibadan South-west Local Government Area (LGA), Oyo State, Nigeria

## Abstract

**Introduction:** Patient satisfaction plays a critical role in healthcare service delivery. Despite growing studies, patient satisfaction levels are still widely under-reported in Nigeria. This study assessed patient satisfaction among patients in primary and secondary healthcare facilities in Ibadan Southwest local government area of Oyo State, Nigeria.

**Method and Materials:** In this cross-sectional study, investigators collected data using the Short-form Patient Satisfaction Questionnaire (PSQ-18). The data was analyzed using the Statistical Package for the Social Sciences (SPSS) software with descriptive statistics and One-way analysis of variance (ANOVA).

**Results:** A total of 387 patients participated in the study, and results showed that overall satisfaction among patients was 67.9%, where Communication (84.5%) and Financial Aspect (54.9%) recorded the highest and lowest percentage of positive responses respectively.

**Conclusion:** Patient satisfaction is a growing subject in Nigeria. It has a significant influence on healthcare service delivery. Stakeholders, policymakers, and managers should employ policies, strategies, and programs to improve the quality of healthcare service delivery across Nigeria.

## Introduction

Over the years, patient satisfaction has become a crucial factor used to determine the performance of healthcare facilities. It is described as a reflection of how content a patient is with their overall experience at their health care service provider.^1^

The origins of patient satisfaction in health care can be traced back to Press Ganey in 1985 when patient satisfaction surveys gained popularity among healthcare institutions that understood the benefit of monitoring patient satisfaction and comparing it to other institutions in a comparable industry.^2^

Patient satisfaction has taken prominence in efforts to enhance healthcare quality worldwide, as there is a growing recognition of the significance of patient-centered care and its influence on general health outcomes.^3^ The development of patient engagement strategies such as E-health tools to improve access to care, innovative communication strategies, and patient satisfaction survey tools are examples of the high level of commitment shown towards improving patient satisfaction.^4,5,6^

Several factors influence patients’ satisfaction during care, including interpersonal communication with health care professionals, technical quality of care, cost of care, time spent with health care professionals, and waiting time.^7^ Recent studies have highlighted relationships between patient satisfaction and patients’ attitudes towards care.^8^ Ensuring that patients are satisfied, well understood, and respected during care is critical, which can lead to a positive attitude towards care. For instance, an unsatisfied patient might seek an alternative form of care if their experience at a healthcare facility was unpleasant.^9^ Furthermore, communication is a reliable predictor of patients’ attitudes towards care, where patients’ desire to accept follow-up visits significantly correlated with their readiness to communicate with healthcare professionals.^10^

In addition, studies have linked long waiting times with dissatisfaction of patients receiving care in health care facilities as patients frequently had specific durations of stay at a medical facility and exceeding this would contradict their expectations.^11,12^

Achieving satisfaction among patients regarding care can be challenging, especially when patients’ expectations do not align with the healthcare institutions’ goals.^13^

Patient feedback, obtained through questionnaires or interviews, is crucial for assessing patient satisfaction in healthcare. It provides insights into staff communication, waiting times, and amenities, enabling practitioners to improve patient experience.^14^

Nigeria and other low- and middle-income countries face unique difficulties in providing healthcare.^15^ Complexities in improving patient satisfaction include overcrowding in medical facilities, unequal allocation of resources, and issues with communication.^16^ Analyzing the healthcare system in Nigeria offers essential insights into the complexities of patient satisfaction in a continuously changing healthcare environment.

The objective of this study was to assess patient satisfaction among patients in primary and secondary healthcare facilities in Ibadan South-west local government area (LGA) of Oyo State, Nigeria. This study was part of a more extensive study that assessed patient safety culture among healthcare professionals and patient satisfaction in Ibadan South-west LGA of Oyo State, Nigeria.

## Methods

### Study Design and Setting

This descriptive cross-sectional multicenter study was conducted in Ibadan southwest LGA of Oyo State, Nigeria. Ibadan South-west LGA is an urban area of Ibadan city with an estimated population of 397,700 individuals, a landmass of 25.13 km2, a population density of 15,828 km2, and an annual population density of 3.5% from the year 2006 to 2016.^17^ With a population index (persons per facility) of 10,150 persons, Ibadan South-west LGA has 43 hospitals with 22 PHCs and 21 secondary hospitals (16 private and five public secondary hospitals).^18^ The selected LGA comprised two of the three-tier levels of healthcare (Primary and Secondary healthcare facilities). The selected study sites included six primary healthcare facilities, three public secondary healthcare facilities, and three private secondary healthcare facilities in Ibadan Southwest LGA, Oyo State, Nigeria. Selected healthcare facilities offered a variation of in-patient and out-patient services.

### Study Population

The population of this study comprised all adult patients who attended the selected study sites at the time of the study.

### Inclusion and Exclusion Criteria

In this study, investigators included English-speaking outpatients as participants who were present at the selected study sites at the time of the visit.

Investigators excluded inpatients and non-English speaking outpatients present at the selected study sites from the study. In addition minors were excluded from the study.

### Sampling and Sample Size Calculation

The researchers employed a multistage sampling technique to gather information from healthcare professionals in the study’s selected locations. In the first stage, investigators purposely selected Ibadan South-west LGA because it had all three tiers of healthcare facilities (primary, public secondary, and private secondary) necessary to carry out this study and an extensive population index (number of people per healthcare facility). Researchers divided healthcare facilities into three categories in the second stage: primary, public secondary, and private secondary. In the third phase, researchers randomly selected primary, private, and public healthcare facilities participating in the study using a balloting procedure.

Using Cochran’s sample size formula (z^2^pq / e^2^ where q = 1-p), researchers selected 385 patients from the population of 3,565,108 individuals in Ibadan to participate in the patient satisfaction survey. Investigators calculated the sample size with a confidence level of 95% with α of 0.05 and a 5% Margin of Error.

### Data Collection and Instrument

Over the years, several patient satisfaction assessment tools have been designed to evaluate patient experience for healthcare service improvement.

In this study, Investigators obtained data from participants using the Short-form Patient Satisfaction Questionnaire (PSQ-18). The survey tool was condensed from the original PSQ questionnaire developed by Hays & Marshall to assess patient satisfaction concisely. A study conducted in India found the PSQ-18 valid and reliable, with internal consistency across subscales varying from 0.72 to 0.93.^19^

The PSQ-18 yields distinct scores for the following seven sub-scales: Financial Aspects, Time Spent with the doctor General Satisfaction, Technical Quality, Interpersonal Manner, Communication, Accessibility, and Convenience.

The researchers in this study obtained data from participants from 4^th^ August 2022 to 1^st^ November 2022 from the selected study sites.

### Statistical Analysis

Investigators utilized the Statistical Package for the Social Sciences (SPSS) software to determine whether the data had a normal distribution. They added the frequency proportions for strongly agree and agree (for positive statements) and strongly disagree and disagree (for negative statements) to calculate the Percentage of Positive Responses (PPR). Negative statements like “I have to pay more for medical care than I can afford” had a reversed response (5 = Strongly Disagree, 4 = Disagree, 3 = Neutral, 2 = Agree, and 1 = Strongly Agree). Positive statements like “My doctors treat me in a friendly and courteous manner” responded positively (5 = Strongly Agree, 4 = Agree, 3 = Neutral, 2 = Disagree, and 1 = Strongly disagree). Investigators calculated the PPR for each subscale of patient satisfaction by adding the frequency proportions for strongly agree and agree (positive statements) and strongly disagree and disagree (negative statements).

Additionally, researchers utilized a one-way analysis of variance (ANOVA) to compare the mean scores of the three institutions. All underlying assumptions of ANOVA were satisfied by this study.

### Ethical Considerations

The Oyo State Research Ethics Review Committee, Ministry of Health Secretariat, Ibadan AD 13/479/4482 approved ethical considerations for this study. The researchers requested permission from the selected healthcare facilities to carry out this study. This study posed no risk or harm to its participants. Participants gave their written and verbal agreement after being informed about the study’s purpose and specifics. Participants were also assured that their answers would remain confidential and anonymous.

Investigators informed participants that participation was voluntary and that they could refuse to participate in the study. In addition, there were no repercussions for individuals who chose to end the study early or continue it at any point. Furthermore, investigators told participants that the study results would be disseminated in aggregate form to academic journals, policymakers, and healthcare managers without identifying their responses.

## Results

Investigators distributed 400 copies of the short-form patient satisfaction questionnaire (PSQ-18) to patients across the selected healthcare facilities. Participants completed and returned 387 questionnaires with a 96.8% response rate.

### Perceptions of Patient Satisfaction among Patients Attending Hospitals in Ibadan South-west Local Government

Table 1. shows perceptions of patient satisfaction among patients attending hospitals in the Ibadan south local government area. Interpersonal manner (75.7%), and communication (84.5%) recorded the highest positive responses. Other subscales of patient satisfaction recorded a percentage of positive responses below 70%. The financial aspect (54.9%) recorded the lowest percentage of positive responses.

**Table 1.**
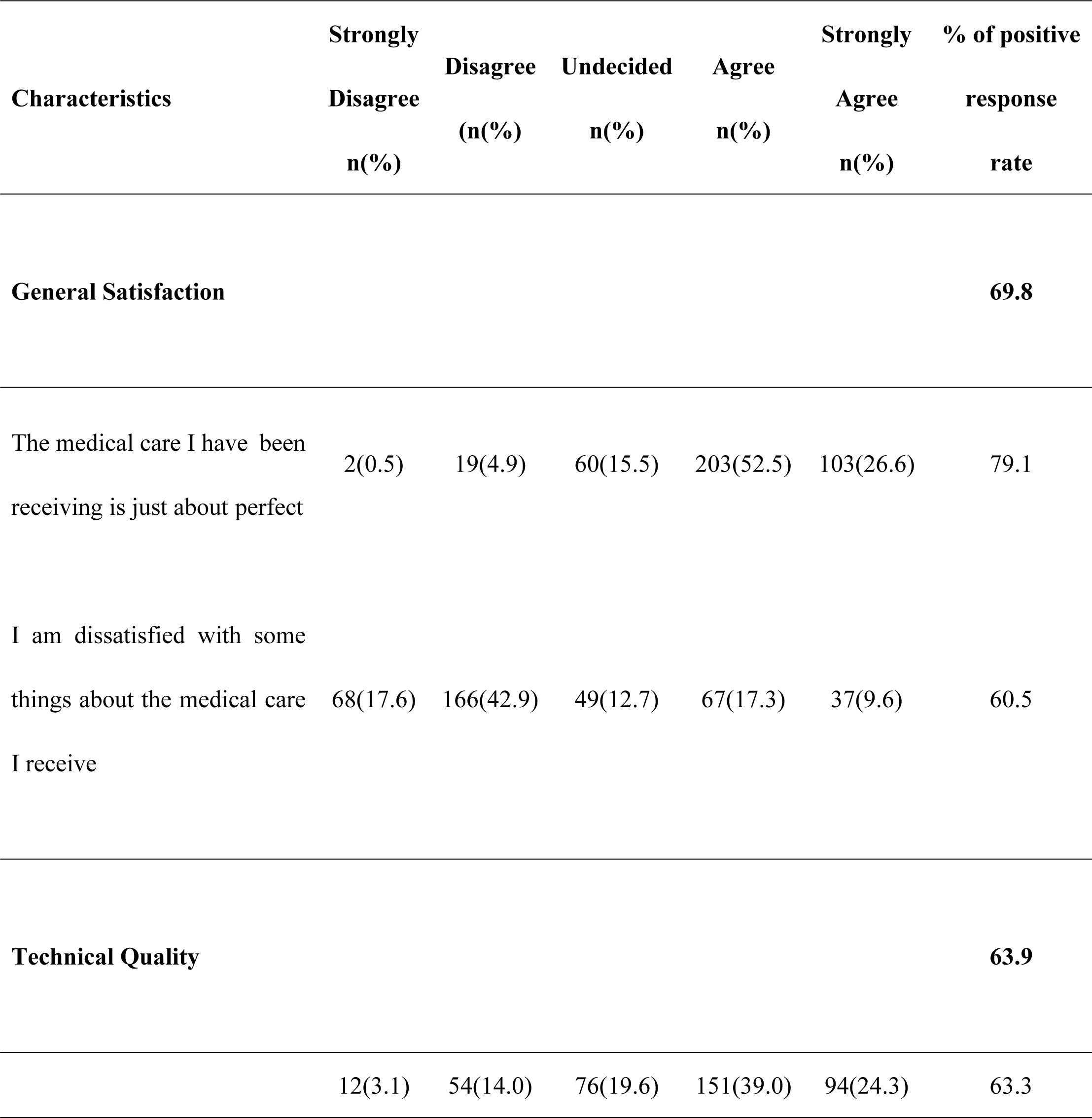
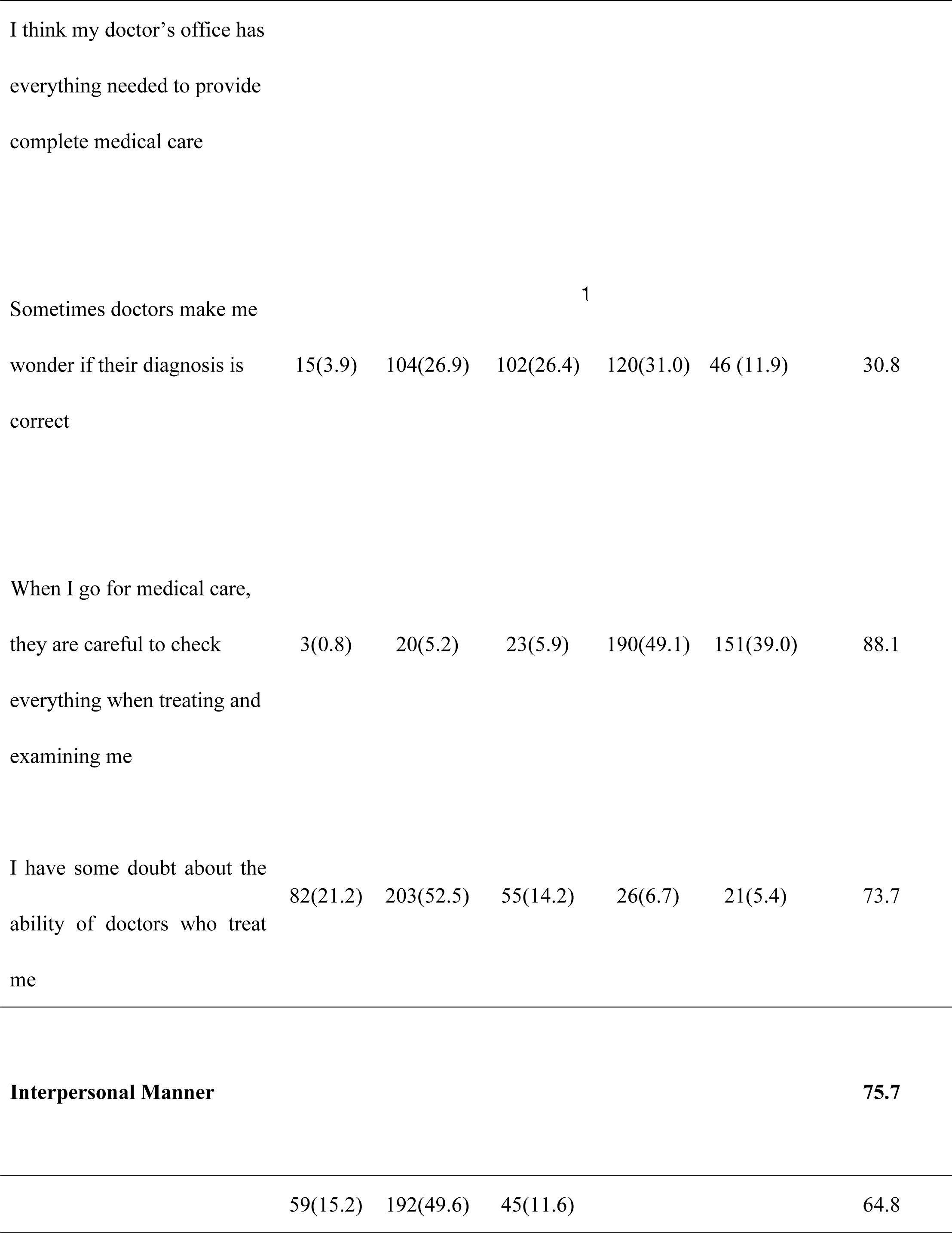
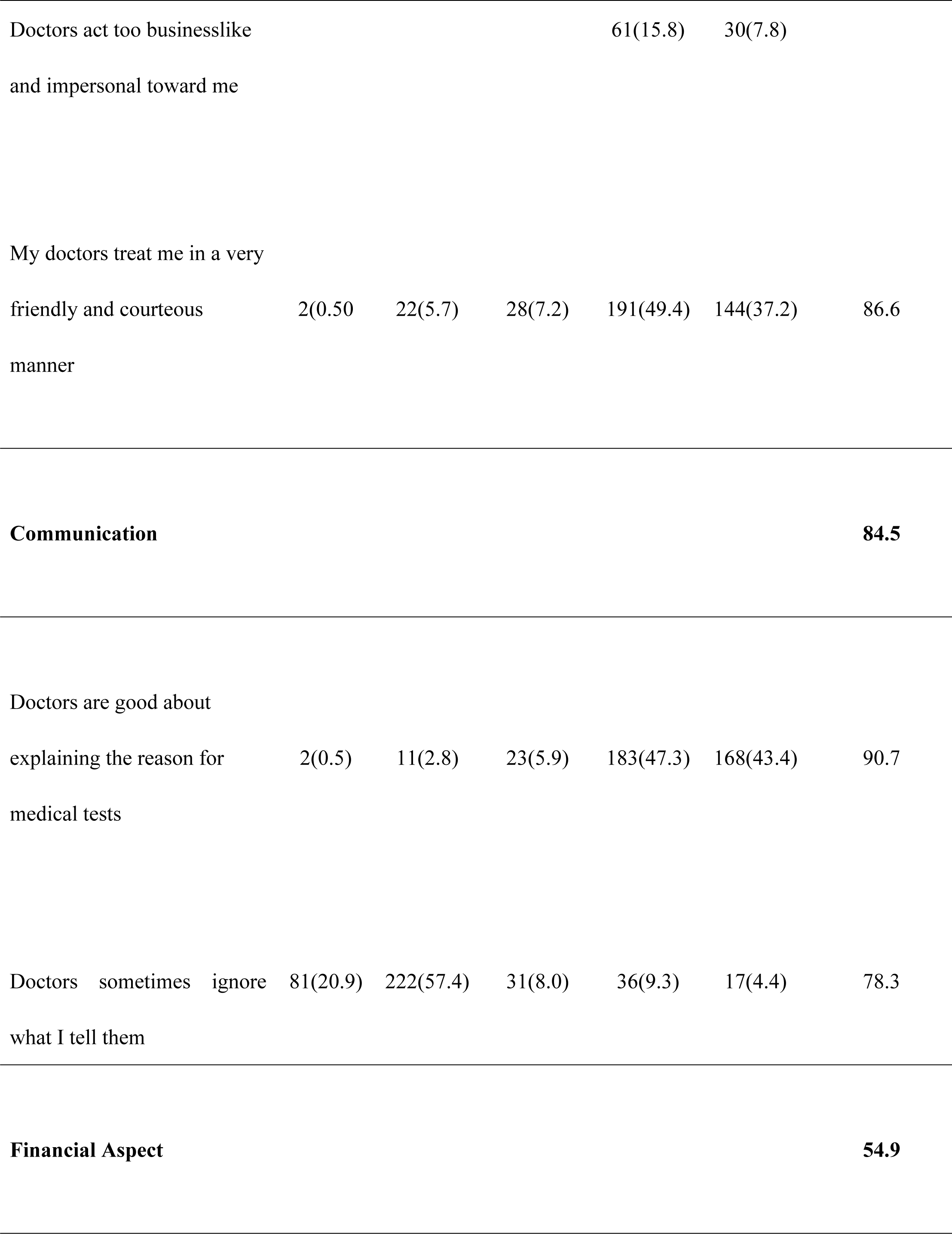
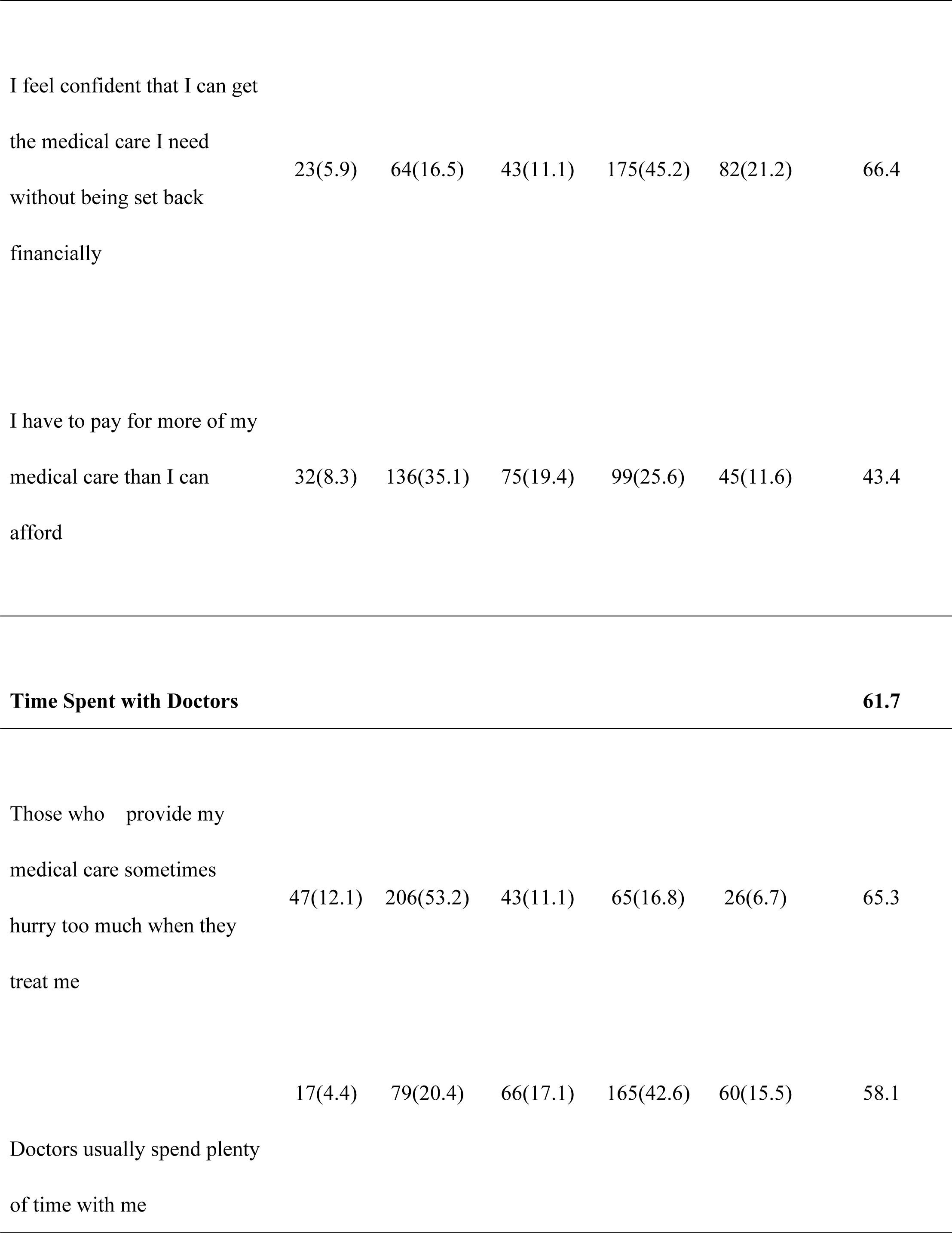
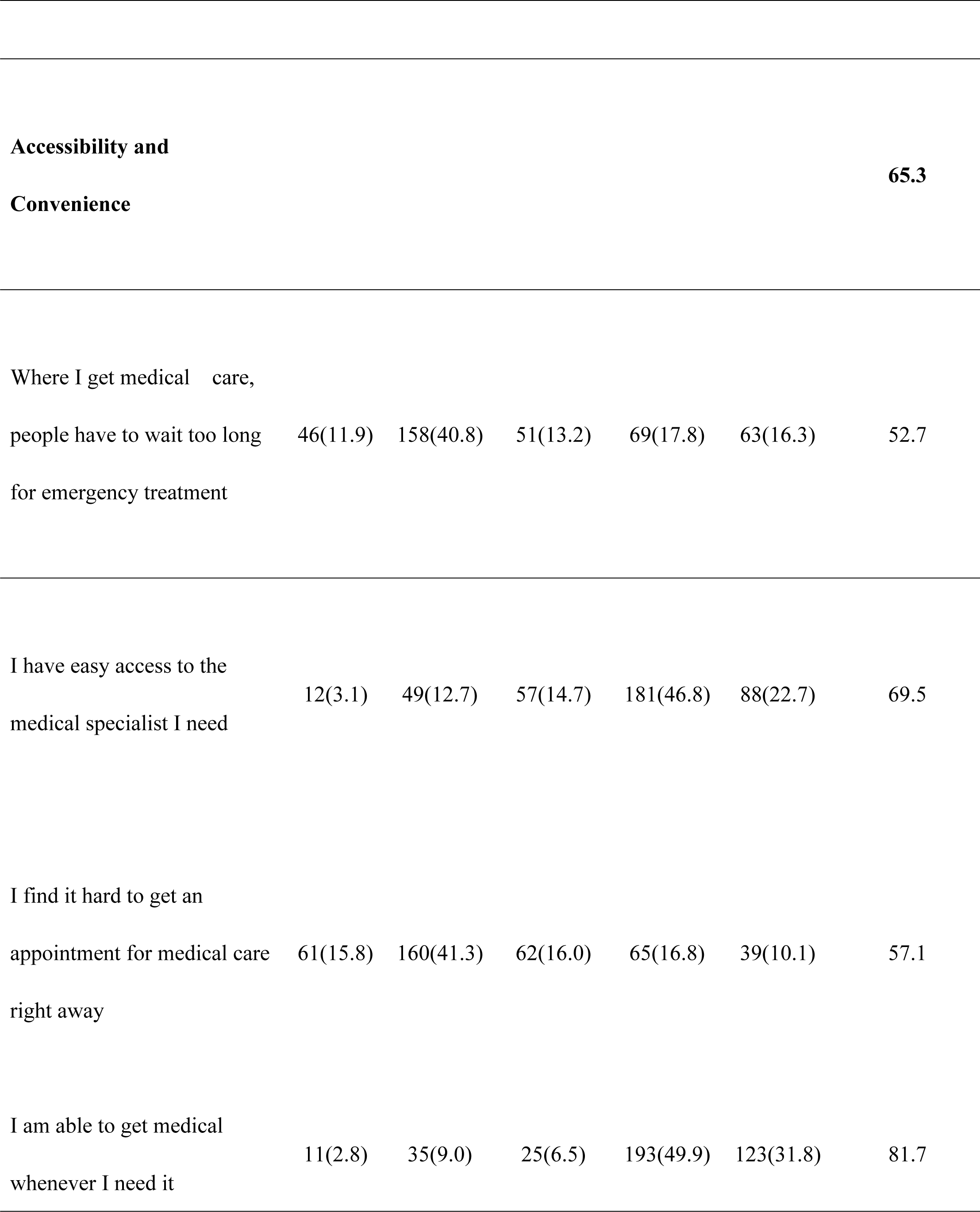
Frequency Distribution on Perceptions of Patient Satisfaction among Patients Attending Hospitals in Ibadan South-west Local Government.

Table 2. The findings of this study also showed that there was no significant difference between the perception of patient satisfaction in the different levels of healthcare facilities in Ibadan South-west LGA.

**Table 2.**
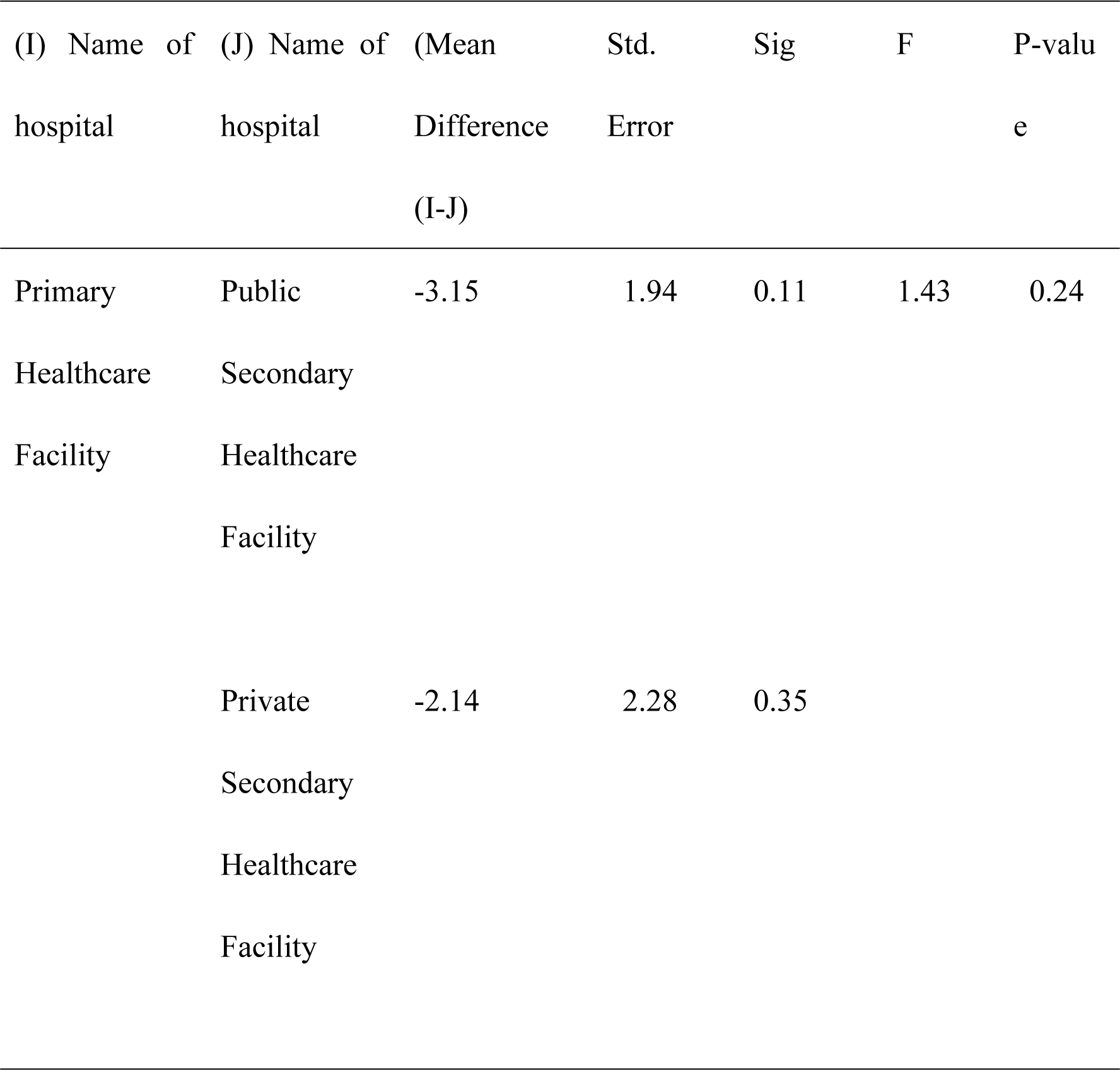
Mean Differences in Patient Satisfaction in the Different Levels of Healthcare Facilities.

## Discussion

This study assessed patient satisfaction in Ibadan South-west LGA, Oyo State, Nigeria. The high response rate from this study corresponded with the response rate from another study conducted in Saudi Arabia.^20^ Among the subscales of the PSQ-18, the financial aspect (54.9%) recorded the lowest Percentage of Positive Responses (PPR). This result was similar to a multicenter study conducted in India, where the financial aspect recorded the lowest satisfaction score.^21^ Also in this study’s subscale analysis, at least 22.4% of patients answered that they could not get healthcare services without being set back financially, and 37.2% paid more for medical care than they could afford. These findings correlated with studies that showed that the cost of medical care in Nigerian healthcare facilities is high.^22,23^

Among PSQ-18 subscales, communication (84.5%) recorded the highest PPR. About 90.7% of participants responded that doctors were good about explaining reasons for medical tests, and 78.3% disagreed that doctors sometimes ignored their views. This result was similar to findings from another study conducted in the United States, where communication recorded a highly positive response. Effective communication extends beyond the exchange of medical information. It encompasses addressing the emotional needs of patients. In addition, healthcare professionals and patients must work together to make decisions that align with the patient’s values, preferences, and goals. By involving patients in the decision-making process, healthcare providers can improve patient satisfaction and contribute to better treatment adherence and outcomes.

Other subscales such as general satisfaction, accessibility, convenience, time spent with doctors, and technical quality, recorded positive responses below 70%, indicating a need for improvement. For instance, the results showed that 26.9% of participants were dissatisfied with some things about the medical care they received, 23.6% responded that doctors acted too businesslike and impersonal towards them, and 34.1% of participants responded that where they received care, people had to wait too long for emergency treatment. These satisfaction levels are also seen in a study conducted in India, where the average level of satisfaction across all subscales was 55.3%.^24^

Regarding the mean differences in patient satisfaction across the different levels of healthcare facilities, this study showed no significant difference in patient satisfaction levels across study sites. While the lack of statistical differences may point towards uniformity in patient satisfaction levels, it is equally essential to consider the possibility of common challenges that may be pervasive across all levels of healthcare facilities.

The study omits sociodemographic data collection to focus on patient satisfaction within a specific local government area, ensuring relevance and applicability to the targeted community. This simplified data collection processes and reduced participant burden, allowing for a more focused analysis of patient satisfaction metrics. However, this omission has limitations, such as compromised generalizability and potential bias in interpretation. This incomplete understanding of patient satisfaction dynamics may hinder efforts to address healthcare disparities and limit the scope of recommendations for service enhancement.

## Conclusion

This study assessed patient satisfaction in Ibadan South-west LGA, Oyo State, Nigeria. The results of this study showed the need for improvement in patient satisfaction levels. To progress toward a more equitable and patient-centered approach, healthcare professionals should address the high cost of care and its impact on patients’ financial aspects. This complex issue calls for cooperation, transparency, and systemic changes. Overall, healthcare managers, policymakers, and stakeholders should employ a proactive and holistic approach to increase patient satisfaction, lower financial barriers, and guarantee that healthcare remains a fundamental right rather than a financial burden.

To satisfy each patient’s unique needs, preferences, and values, healthcare providers should consider customizing their treatments. Healthcare practitioners can foster a more compassionate and individualized experience that positively resonates with patients by including them in decision-making, respecting their autonomy, and considering their emotional well-being.

## Data Availability

The data associated with this study are available at the following Figshare repository: 10.6084/m9.figshare.25295449.

## Acknowledgment

The authors would like to thank the management of the selected study sites and participants.

## Declaration of Conflicting Interests

The author(s) declared no potential conflicts of interest concerning the research, authorship, and/or publication of this article.

## Ethical Approval

The Oyo State Research Ethics Review Committee, Ministry of Health Secretariat, Ibadan AD 13/479/4482 granted this study ethical considerations.

## Funding

The author(s) received no financial support for this article’s research, authorship, and/or publication.

## Notes

### Competing Interest Statement

The authors have declared no competing interest.

